# SARS-CoV-2 Introductions into Lao PDR Revealed by Genomic Surveillance, 2021–2024

**DOI:** 10.64898/2026.04.09.26349480

**Authors:** Siribun Panapruksachat, Cécile Troupin, Manila Souksavanh, Chantisa Keeratipusana, Manivanh Vongsouvath, Susath Vongphachanh, Malavanh Vongsouvath, Koukeo Phommasone, Somphavanh Somlor, Matthew T. Robinson, Thanat Chookajorn, Theerarat Kochakarn, Nicholas PJ Day, Mayfong Mayxay, Andrew G. Letizia, Audrey Dubot-Pérès, Elizabeth A Ashley, Philippe Buchy, Phonepadith Xangsayarath, Elizabeth M. Batty

## Abstract

We used 2492 whole genome sequences from Laos to investigate the molecular epidemiology of SARS-CoV-2 from 2021 through 2024, covering the major waves of COVID-19 disease in Laos including time periods of travel restrictions and after relaxation of travel across international borders. We identify successive waves of COVID-19 caused by shifts in the dominant lineage, beginning with the Alpha variant in April 2021 and continuing through the Delta and Omicron variants. We quantify a shift from a small number of viral introductions responsible for widespread transmission in early waves to a larger number of introductions for each variant after travel restrictions were lifted, and identify potential routes of introduction into the country. Our study underscores the importance of genomic surveillance to public health responses to characterize viral transmission dynamics during pandemics.

## Introduction

Severe acute respiratory syndrome coronavirus 2 (SARS-CoV-2) was first reported in Wuhan, China in December 2019^1^ and COVID-19 was declared a pandemic on March 11th 2020. Spreading globally over the past 5 years with multiple genetic lineages, including the Alpha, Beta, Delta, and Omicron variants of concern, this respiratory virus continues to be a source of morbidity and mortality^2^. Ongoing sequencing to identify novel lineages is necessary to investigate epidemiology and aid in the development of preventive measures, including vaccines targeting contemporary strains^3,4^.

Laos is a landlocked country sharing borders with Cambodia, Thailand, Myanmar, China, and Vietnam. Laos has only one major international airport and 40% of the 4.8 million international arrivals in 2019 arrived by air, with the rest arriving through 19 land borders^5^. The first cases of SARS-CoV-2 infection in Laos were detected on March 24th 2020^6^. However, despite the early circulation of the virus in Laos, the number of cases and the seroprevalence remained low throughout 2020^7^. Mass gatherings were limited and entry by land and air was restricted in April 2020 with new arrivals quarantined in hotels and quarantine facilities for two weeks whether arriving by land or air. Other Southeast Asian countries also reported low case numbers and seroprevalence throughout 2020^8,9^, decreasing the likelihood of SARS-CoV-2 importation into Laos through land borders from neighbouring countries. In April 2021, the number of COVID-19 cases increased, leading to further restrictions on social gatherings including the closure of entertainment venues and restricting entry to the country only to persons with a negative RT-PCR test^10^. Vaccination commenced in April 2021, and by January 2022, 49.9% of the population had been fully vaccinated. Entry of fully vaccinated tourists with a negative RT-PCR test on arrival was allowed in January 2022, and international borders fully reopened in May 2022.

Whole genome sequencing emerged as a key tool to track the evolution of SARS-CoV-2 throughout the pandemic. The Pango lineage classification system was developed early in the pandemic^11^ to designate new lineages as a tool for tracking the movement of viruses between locations. The World Health Organization (WHO) subsequently classified particular lineages as Variants of Concern and assigned simple labels (Alpha, Delta, Omicron) for key lineages^12^. We used whole genome sequences of SARS-CoV-2 strains collected in Laos between 2021 and 2024 to investigate the viral genotypes circulating in Laos, a country with limited prior experience of genomic surveillance of epidemic pathogens. As a country with few international airports and border crossings, and which enacted a strict quarantine protocol and relaxation of borders during the time period where there was active genomic surveillance, it serves as an excellent location to evaluate introduction events during and after pandemic mitigation measures. Additionally, these results can help inform the transmission dynamics of a novel virus during border security operations for the US military helping to safeguard our service members while protecting the country from introductory viral events.

## Methods

### Sample collection and sequencing

Samples were collected as part of the public health activities carried out by the Lao-Oxford-Mahosot Hospital-Wellcome Trust Research Unit (LOMWRU), and Institut Pasteur du Laos (IPL). These included specimens collected during routine hospital screening, testing of contact cases obtained during epidemiological investigations conducted by provincial or national health authorities, screening of air passengers, and patients enrolled in a case-based surveillance study^13^.

For LOMWRU samples, nasopharyngeal and oropharyngeal swabs were collected and all samples that tested positive for SARS-CoV-2 by quantitative reverse transcription polymerase chain reaction (RT-qPCR) with a Ct value below 28 were selected for sequencing (Supplementary Table 1). RNA was extracted using QIAamp Viral RNA Mini kit (Qiagen) or Nucleic Acid Extraction Rapid kit (Bioperfectus) according to the manufacturer’s instructions. RNA was reverse transcribed using LunaScript RT (NEB) and SARS-CoV-2 amplicons were generated using the Oxford Nanopore Technologies (ONT) Midnight primer set from Midnight RT PCR Expansion kit (EXP-MRT001). ONT sequencing libraries were generated using the Rapid Barcoding Kit (SQK-RBK110.96) according to the manufacturer’s instructions and sequencing on the ONT MinION Mk1B sequencer. Up to 96 samples were batched on one R9.4 flow cell. Raw sequence data were basecalled using Guppy in high-accuracy mode.

For IPL samples, viral RNA was extracted from nasopharyngeal and/or oropharyngeal swabs by NucleoSpin RNA virus kit (Macherey-Nagel), NucleoSpin 8 virus kit (Macherey-Nagel) or Nucleic Acid Extraction Rapid kit (Bioperfectus) according to the manufacturer’s instructions. Extracted RNAs were then screened for the detection of SARS-CoV-2 genome by RT-qPCR following the Charité-Berlin protocol^14^. Samples with Ct values below 30 were selected for sequencing. IPL used the highly multiplexed PCR amplicon approach developed by Quick *et al.*^15^ with the ARTIC Network multiplex PCR primers set v4.1 and v.5.3.2 primers from IDT. Briefly, viral RNA was reverse-transcribed using LunaScript RT SuperMix (New England Biolabs) and SARS-CoV-2 amplicons were obtained using the Q5 HS Master Mix (New England Biolabs). Samples were multiplexed using Oxford Nanopore rapid barcode kit (SQK-RBK110.96) and run in batches of 4-24 samples on R9 flow cells using the MinION Mk1B sequencer. Basecalling and demultiplexing were performed by MinKNOW software using Guppy or Dorado in super-accurate mode.

### Ethical approval

The protocol for the case-based surveillance study was approved by the Lao National Ethics Committee for Health Research and the Oxford Tropical Research Ethics Committee (ref 53-20). No ethical approval was required for sequencing performed in the framework of national surveillance requested by the Lao Ministry of Health.

### Bioinformatic analysis

Basecalled sequence data was analysed using the ONT wf-artic workflow^16^ as implemented in EPI2ME to generate consensus genomes for the LOMWRU samples or using the COVID-19 EDGE pipeline^17^ for the IPL samples. Genome sequences from IPL were uploaded to GISAID, and genome sequences from LOMWRU were uploaded to both GISAID and GenBank, except for those sequences containing frameshift mutations which were rejected by GenBank submission (Table S1). PANGO lineages were determined using Nextclade v3.11.0^18^. Individual lineages were grouped into the Variants of Concern and Variants of pango-collapse v0.8.2^19^. A phylogenetic tree of all public SARS-CoV-2 strains was downloaded from https://hgdownload.gi.ucsc.edu/goldenPath/wuhCor1/UShER_SARS-CoV-2/ on 1^st^ October 2024 and the Laos samples not already included in this tree were placed using UShER^20^. The matUtils package was used to analyse the tree and determine introductions of SARS-CoV-2 into Laos using the matUtils introduce function^21^. matUtils extract was used to extract subtrees for each variant with the most closely related samples from the global tree. The public UShER tool available at https://genome.ucsc.edu/cgi-bin/hgPhyloPlace was used to find the most similar genomes to individual strains available in GenBank or GISAID. To identify representative strains for each introduction event, the sample with the earliest collection date was selected. Introduction events with multiple strains sharing the same earliest collection date but originating from different provinces were excluded to avoid ambiguity. The dataset was then categorized into two groups relative to the implementation of the lockdown on May 9, 2022: During Lockdown and After Lockdown.

For each introduction within both groups, we initially extracted 200 closely related strains using the extract function from matUtils. We then iteratively expanded the subtree until the inferred origin traced back to the closest non-Lao node, representing the likely country of origin. Ancestral state reconstruction was performed using PastML^22^ with the DELTRAN method, and polytomies were resolved. The most likely geographic origin of internal nodes was inferred. A dataset with number of reported COVID-19 cases from Laos was downloaded from Our World In Data^23^. Phylogenetic trees were visualised using the ggtree^24^ and ggtreeExtra^25^ packages in R.

## Results

We analysed the complete genome sequences of 2492 SARS-CoV-2 strains collected between April 2021 and September 2024 and sequenced between April 2022 and September 2024. The strains were collected from 17 of the 18 provinces in Laos. The strains obtained from Vientiane Province and Vientiane Capital represented 57.71% of all the samples included in the study (Figure 1). The number of strains sequenced per week was proportional to the number of cases detected per week for much of the time period studied, except for the period November 2021- February 2022. During these 3 months, the number of cases was relatively high while the number of strains sequenced remained low. Additionally, after April 2023, the number of strains sequenced by our laboratories was higher than the number of cases reported to Our World in Data, likely due to under-reporting of RT-qPCR testing or due to initial diagnosis by rapid diagnostic tests (RDT) (Figure 2). The overall percentage of cases sequenced during the period covered by this study is 1.18%, which is below the suggested proportion of 5% needed to detect lineages at a prevalence of 0.1 to 1.0%^19^. The percentage of cases sequenced per week is shown in Figure 2C. The red dotted line represents the suggested proportion of 5% needed to detect lineages at a prevalence of 0.1 to 1.0%^26^. As sequencing activity for SARS-CoV-2 virus was implemented in LOMWRU in May 2022 and at IPL in November 2022, many strains were actually sequenced retrospectively, with a long delay between the sample collection date and the sequencing date (median value 268 days), although this interval decreased throughout the pandemic (Figure S1). The mean delay after November 2022, when both sites had implemented sequencing workflows, was 64 days.

**Figure 1.**
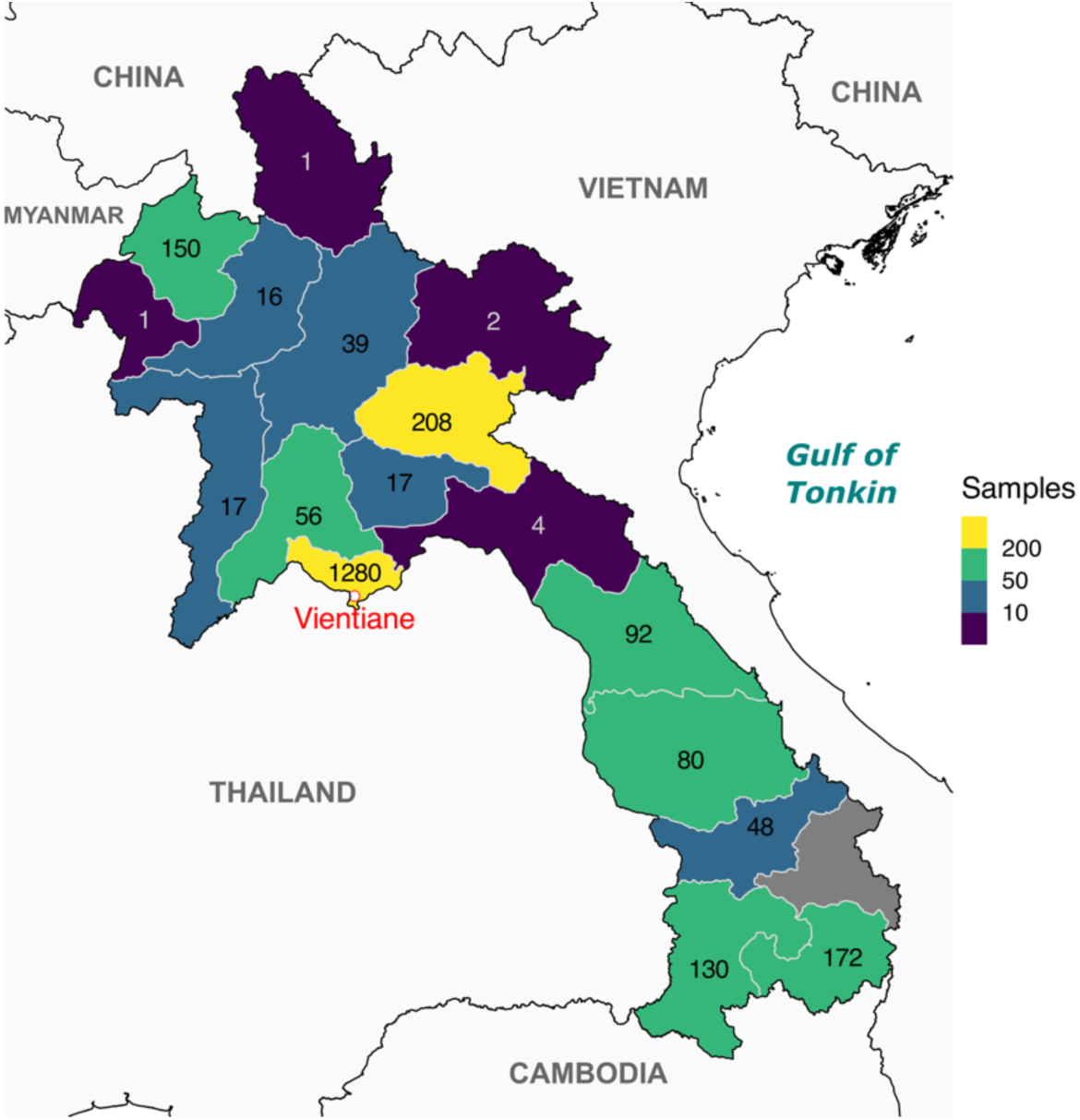
Map showing the repartition by province of the 2492 SARS-CoV-2 strains included in the study. Of note, no samples were obtained from Sekong Province. The active case-based surveillance study took place in Luang Namtha, Xiangkhouang, Salavan and Attapeu provinces.

**Figure 2.**
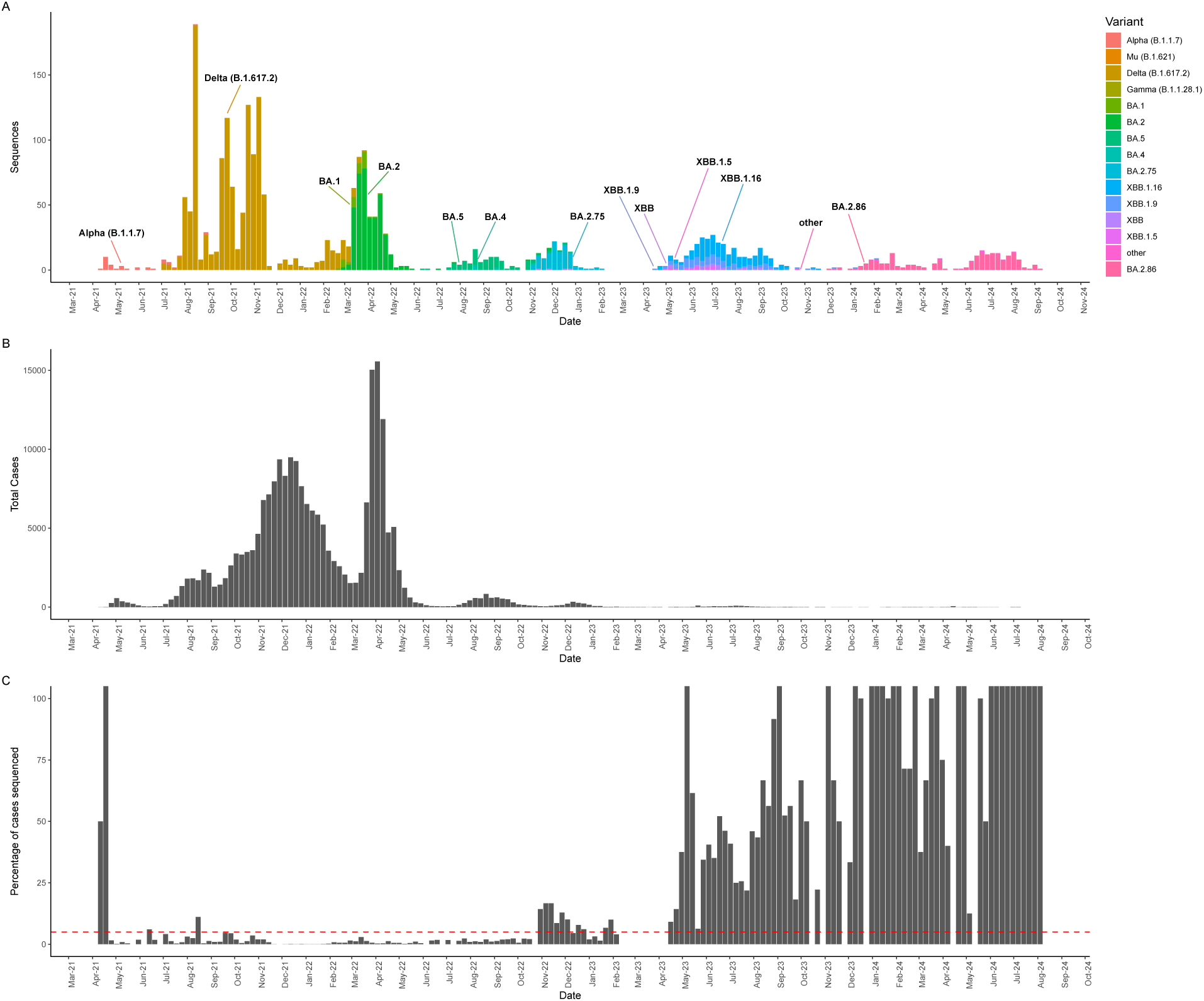
A - A plot of number of strains sequenced each week in this project, coloured by variant. B – Number of COVID-19 cases reported per week in Our World in Data reports15. C - Percentage of weekly COVID-19 cases that were sequenced. Some weeks show percentages exceeding 100 percent due to under-ascertainment of the COVID-19 cases, and the Y-axis has been truncated at 100

The strains sequenced fell into 162 different Pango lineages. For further investigation, we grouped the Pango lineages into broader Variants of Concern based on the parent Pango lineage (Table 1 and Figure 2).

**Table 1.** The number of strains, dates of first and last detection in Laos, and province where the strain was first detected for each variant analysed. Multiple locations of first detection are given when the variant was detected in multiple provinces on the same day.

| Variant | Number of strains | First detected | Last detected | Location of first detection |
| --- | --- | --- | --- | --- |
| Alpha (B.1.1.7) | 43 | 11/04/2021 | 31/08/2021 | Vientiane Capital |
| BA.1 | 42 | 25/02/2022 | 28/04/2022 | Unknown |
| BA.2 | 399 | 01/03/2022 | 14/12/2022 | Xiangkhouang |
| BA.2.75 | 113 | 08/11/2022 | 28/04/2023 | Luang Namtha |
| BA.2.86 | 199 | 08/12/2023 | 09/09/2024 | Vientiane Capital |
| BA.4 | 8 | 11/08/2022 | 07/11/2022 | Vientiane Capital |
| BA.5 | 97 | 15/06/2022 | 26/12/2022 | Vientiane Capital |
| Delta (B.1.617.2) | 1245 | 04/07/2021 | 28/03/2022 | Champasak, Khammouan, Savannakhet |
| Gamma (B.1.1.28.1) | 1 | 24/11/2021 | 24/11/2021 | Xiangkhouang |
| Mu (B.1.621) | 1 | 06/2021 | 06/2021 | Vientiane Capital |
| XBB | 32 | 16/11/2022 | 21/09/2023 | Vientiane Capital |
| XBB.1.16 | 190 | 27/04/2023 | 20/11/2023 | Vientiane Capital |
| XBB.1.5 | 12 | 03/05/2023 | 06/07/2023 | Savannakhet |
| XBB.1.9 | 95 | 21/04/2023 | 04/02/2024 | Vientiane Capital |
| Other variant | 15 | 13/05/2023 | 09/08/2024 |  |

One early strain (EPI_ISL_19276765) detected in Vientiane Capital in April 2020 belonged to the B.1.1 variant, which differed by only 9 mutations from the reference Wuhan strain. The first variant of SARS-CoV-2 to cause a substantial number of infections in Laos was the Alpha variant (Pango lineage B.1.1.7), first detected in April 2021. The earliest collected strain, IPL-0731 (EPI_ISL_19276767), was obtained in Vientiane Capital and is most closely related (a difference of 2 SNPs) to a set of over 40 identical strains from Germany and Belgium. Five independent introductions of this variant occurred, with four of the introductions leading to 1-3 strains detected only in Laos, while the second introduction led to the largest cluster, comprising 37 strains obtained from 21 April to 31 August 2021. The earliest strain collected from this cluster is IPL-07920 (EPI_ISL_19276772) which is identical to strains from China, Vietnam, the USA, and Thailand (Figure 3).

**Figure 3.**
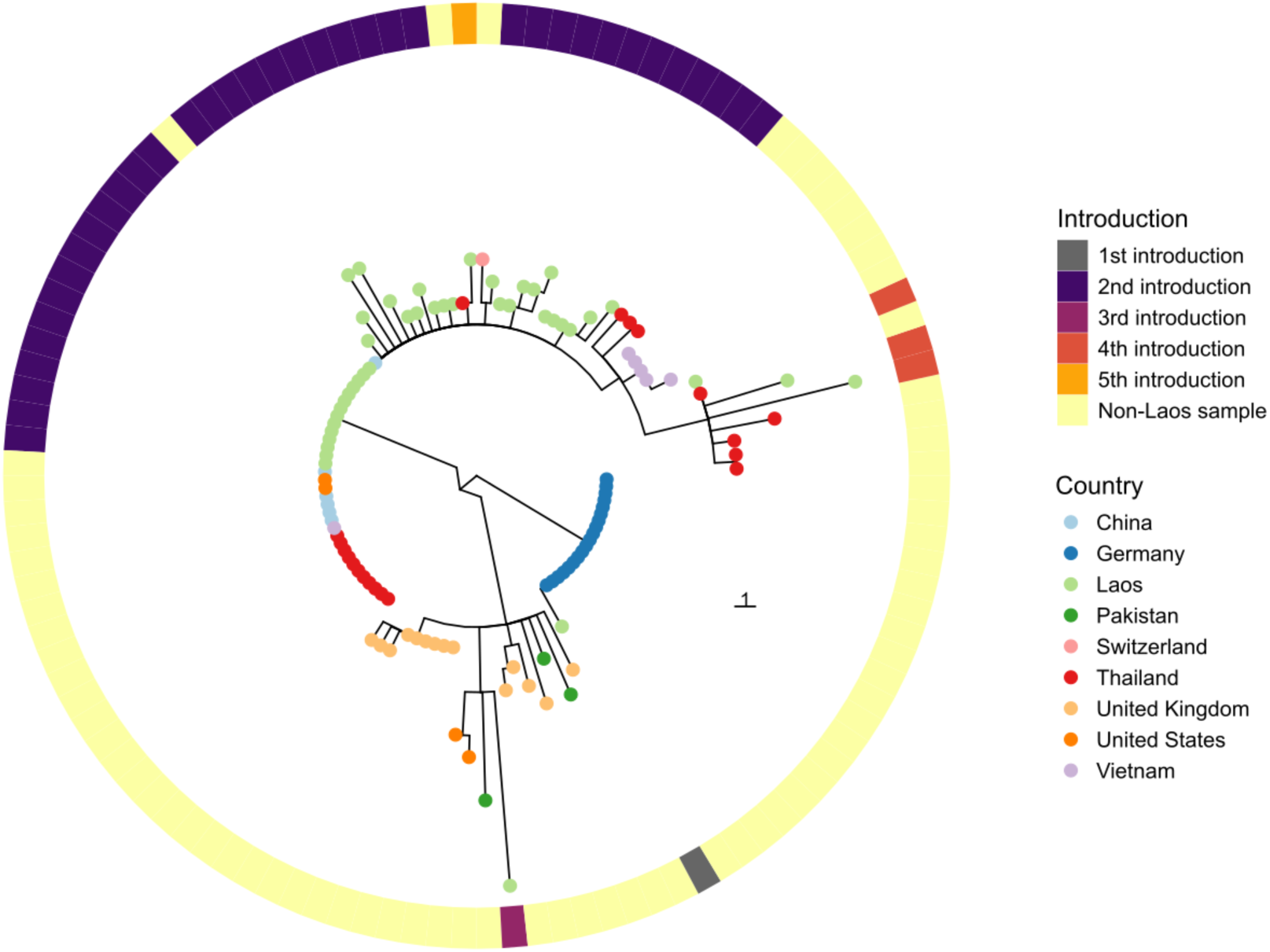
Phylogenetic tree generated using UShER showing the 43 sequenced Alpha variant samples from Laos and the 73 most closely related samples from the global tree. The coloured clades show the number of introductions of the Alpha variant into the country. Coloured tip labels identify the probable country of origin for each strain. The scale bar is measured in number of mutations.

The Delta variant (lineage B.1.617.2 and descendants) was the predominant lineage detected among our strains in Laos, circulating from July 2021 to March 2022. A total of 1245 of the 2492 strains (50%) belong to the Delta variant, and they are predominantly from the AY.1 (633 strains, 51%) and AY.85 (482 strains, 39%) lineages (Figure 4). Analysis of the introduction events showed a single importation of the AY.1 and AY.85 variants into Laos that consecutively led to large clusters and subsequent proliferation of descendant strains. To investigate the transmission chains of AY.1 and AY.85, we identified the most closely related strains from the global public tree using UShER and generated a phylogenetic tree of these strains (Figure 4). The first recorded AY.1 strain was sampled on 28th July 2021 (strain LOMWRU-1433, accession ID EPI_ISL_18001417). The most closely related strains from outside Laos are from Germany and were sampled in early August, after the AY.1 variant was already circulating in Laos (Figure S4). The first recorded AY.85 strain was collected on 4th July 2021 in Savannakhet Province (strain IPL-19375, accession ID EPI_ISL_19276806) and is most closely related to strains from Thailand (Figure S5).

**Figure 4.**
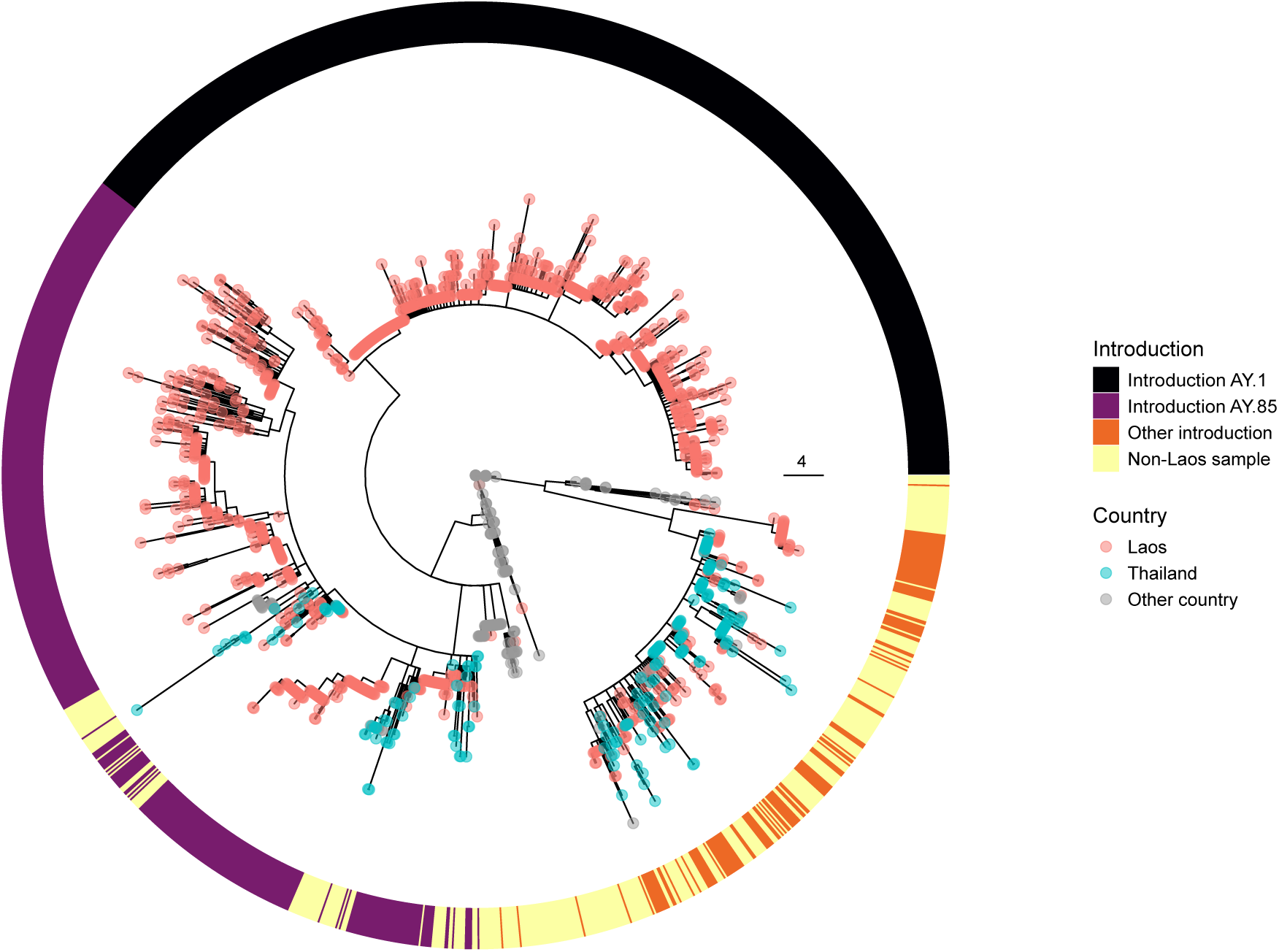
Phylogenetic tree generated using UShER showing the 1245 Delta variant sequences from Laos and the 359 most closely related sequences from the global tree. The purple and black coloured clades show the strains descended from the single introduction of AY.1 and AY.85 into the country, while the orange-coloured clades are Laos strains from other introductions of the Delta variant and yellow strains are closely related strains from outside Laos. Coloured tip labels identify the strains from Laos, Thailand (the second most common probable country of origin), and other countries. The scale bar shows number of mutations.

During the Alpha and Delta waves, two strains identified as the Gamma and Mu variants were detected. The Gamma (P.1) variant of concern strain was collected from a patient in Xiangkhouang province in November 2021 (accession ID EPI_ISL_15656023/OQ028273.1). Strains with identical genome sequences were found in multiple countries including Brazil, Argentina, Germany, the United Kingdom (UK), the United States of America (USA), Uruguay, and Colombia, but no identical strains were found in Asia. The identical strain with the closest collection date to our Laos Gamma strain was collected in late October 2021 in the United Kingdom (OX833583.1), suggesting either a later import of this strain into Laos, or that it was circulating without detection before November 2021 (Figure S2). A single Mu variant (B.1.621) strain was collected in Vientiane Capital in June 2021 (EPI_ISL_19276799). This strain is identical to strains found in the USA, and Colombia (Figure S3). The Mu variant was classed as a Variant of Concern in 2021 and was not previously detected in Laos.

The third wave of COVID-19 in Laos was predominantly caused by BA.1 and BA.2 lineage strains, which are both part of the Omicron variant. Both lineages were first identified only 6 days apart in early 2022 (25th February 2022 and 1st March 2022, respectively). The BA.1 and BA.2 lineages rapidly displaced the final Delta variant strains, which were last seen on 28th March 2022. Despite being introduced at around the same time, the last BA.1 strain was detected on 28th April 2022, while the BA.2 strain comprised the majority of samples until July 2022, with the last BA.2 strain detected in December 2022. The earliest BA.2 strains were identified from Xiangkhouang province, while the earliest BA.1 strains with a known location were identified from Vientiane.

The 42 BA.1 strains form two separate clades which are identified as two separate introductions of BA.1 into Laos (Figure 5). For 399 BA.2 strains, there are 63 separate introductions. The earliest introduction, into Xiangkhouang province, had only 14 descendant strains, while the second introduction, which also occurred in Xiangkhouang province, had 93 descendant strains, forming the largest BA.2 cluster.

**Figure 5.**
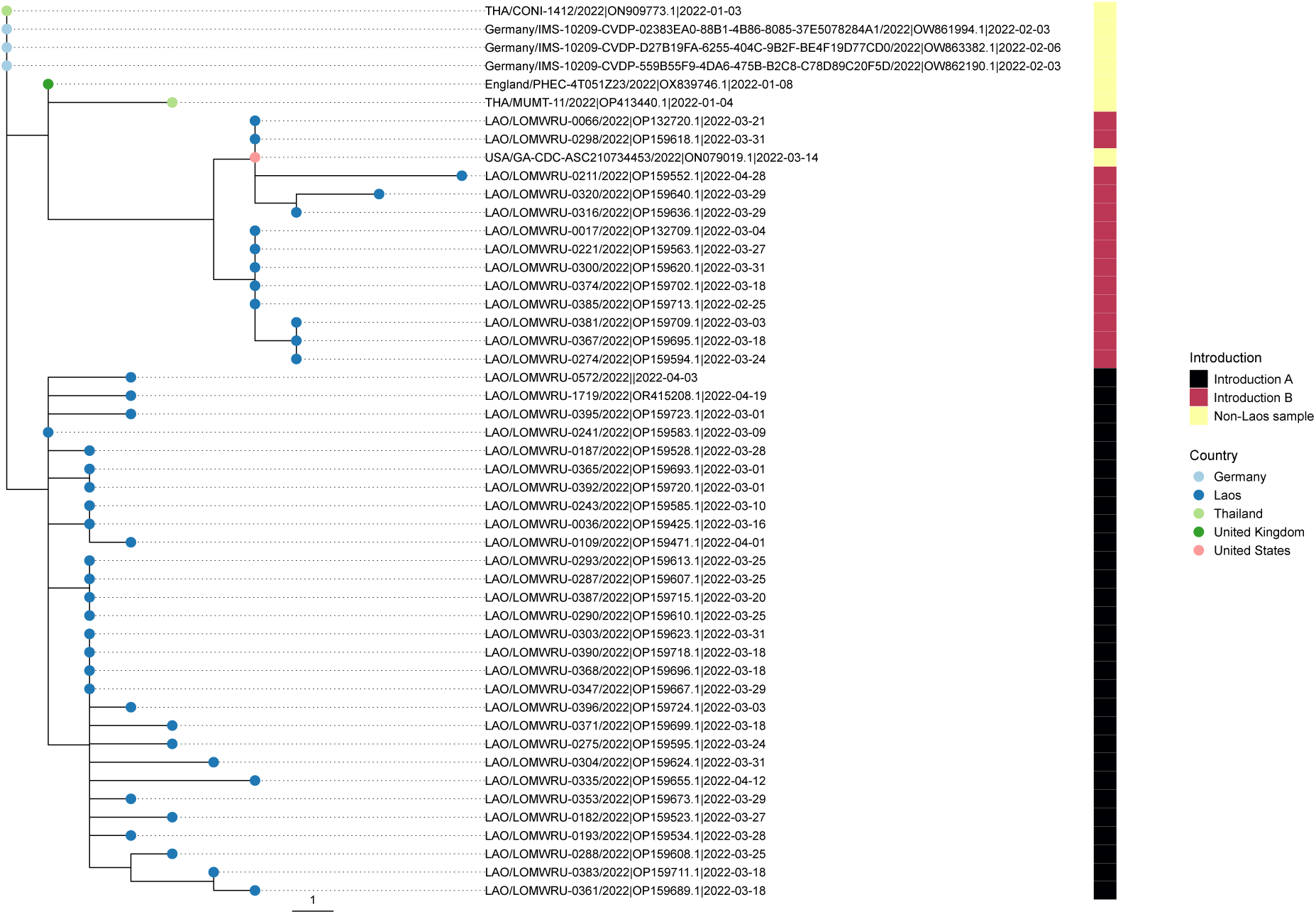
A phylogenetic tree generated using UShER showing all the BA.1 strains sequenced in Laos, with other closely related samples from a global tree. Coloured tip labels identify the country, and the heatmap identifies the two clades descended from separate introductions into the country as well as the non-Laos strains. The scale bar shows the number of mutations.

During the fourth wave of COVID-19, starting from June 2022, the BA.2 variant strains were displaced by BA.5 variants. These strains belonged to the BA.5.2 sub-lineage and were first detected in Vientiane, and then subsequently in other provinces. Only eight BA.4 lineage strains were collected in Laos, suggesting minimal spread of this variant.

Several smaller COVID-19 waves occurred between late 2022 and September 2024 (Figure 1B). During this period, the proportion of cases subjected to sequencing increased significantly, occasionally reaching 100% of all reported cases (Figure 1C). BA.2.75 variant strains were first introduced in early November 2022, and a total of 113 strains representing 17 Pango lineages and corresponding to 37 separate introductions were sequenced. No further strains were sequenced until the introduction of the XBB.1.5 recombinant lineage in May 2023. While the parent XBB lineage was detected in November 2022, widespread transmission did not appear to occur until the introduction of XBB.1.5, which shows increased transmission compared to the parent lineage^27^, followed by XBB.1.16 variants. The dominant lineages from early 2024 were BA.2.86 lineages, including the JN.1, KP.2, and KP.3 lineages.

We used a global tree of public strains to contextualise each Laos SARS-CoV-2 strain and assessed whether each strain represented a novel introduction of the virus from an external country, or a descendant of an existing strain previously introduced into the country. This identified 443 separate introductions from a total of 2492 samples in this study, and assigned each instance as either an introduction or a descendant of an introduced strain. We could then investigate the size of each introduced cluster to determine whether the pattern of introductions into the country changed over time given the changes in non-pharmaceutical interventions, availability of testing, and population infection and vaccination history at different stages of the pandemic. We used the date of May 9^th^ 2022 when all travel restrictions into Laos were lifted^28^ to divide our introductions into two sets, “during-lockdown” and “after-lockdown”, to compare the most likely geographic origin for each introduction into Laos, and the local province for each introduction. A total of 100 and 343 introduction nodes were identified, corresponding to 1537 and 755 strains detected during and after the lockdown periods, respectively. Two introduction nodes from the during-lockdown period and one from the after-lockdown period were excluded due to ambiguity in their spatial origin.

During the lockdown period, 1107 out of 1537 strains (72%) were collected from Vientiane Capital and Vientiane Province, and no strains were collected from Houaphan, Oudomxay, and Phongsali. The after-lockdown group showed a more even geographical distribution of samples, although no strains were collected from Bokeo province during that period (Figure 6A). During both periods, Vientiane Capital had the highest proportion of introductions, accounting for 59% (59/100) and 32.65% (112/343) of the introductions, respectively. After lockdown, five additional provinces had introduction events compared to during the lockdown.

**Figure 6.**
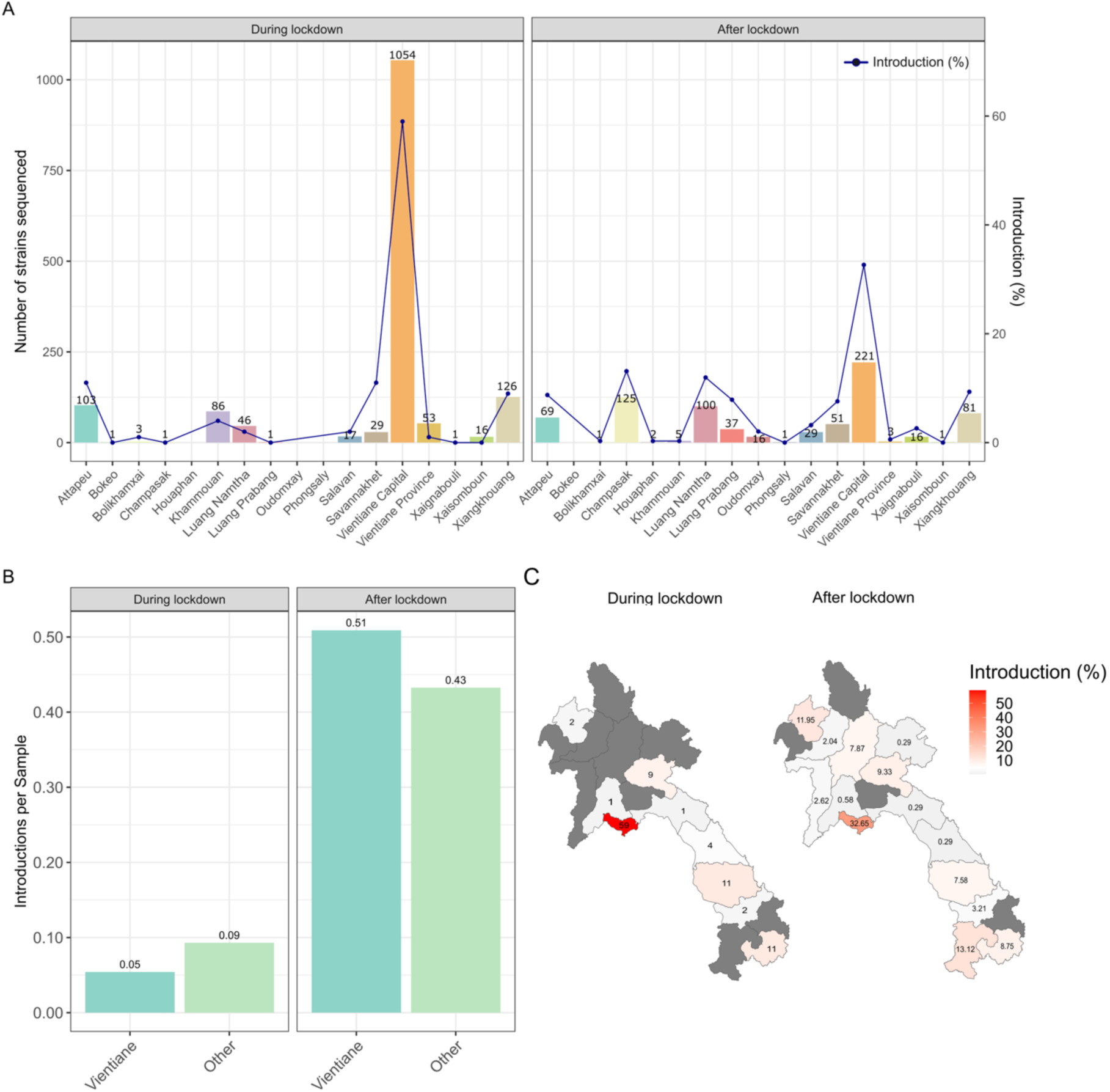
(A) Number of strains sequenced (bar) and percentage of introductions (line) attributed to each province. The exact number of strains sequenced is shown above each bar. (B) Introduction per strain for Vientiane Capital and Vientiane Province compared to other provinces across the two periods. (C) Geographic distribution of percentage of introductions per province during and after the lockdown.

We compared the overall introduction rates between Vientiane (including Vientiane Capital and Vientiane Province) and other provinces during and after lockdown to assess whether travel restrictions affected the location of introductions, as entry to the country during the restriction period would have been limited to air travel, and the main international airport is located in Vientiane. In order to account for the uneven distribution of strain collections between provinces, we normalized the number of introductions by the number of strains to give a measure of introductions per strain, with a higher figure suggesting more introductions with fewer descendant strains, and a lower figure suggesting a smaller number of introductions that gave more descendant strains. In Vientiane Capital and Vientiane Province, the introductions per sample rose from 0.05 during lockdown to 0.51 after lockdown. In contrast, the other provinces showed an increase from 0.09 to 0.43. Interestingly, during the lockdown period, the introductions per sample were higher in the other provinces compared to Vientiane. However, this trend reversed after the lockdown, with Vientiane showing a higher introduction rate than the rest of the country (Figure 6B).

We used PastML to infer a geographic origin for each introduction into Laos. During the lockdown period, the two countries most often inferred as possible origins were Thailand (48/100, 48%) and the United Kingdom (29/100, 29%). In contrast, during the after-lockdown period, the most common possible origins shifted to the United States (199/343, 58%) and China (81/343, 24%). Vientiane Capital, Vientiane Province, Khammouan, Savannakhet and Saravan, all of which share a border with Thailand, had Thailand as the predominant source of introductions during the lockdown period. In the post-lockdown period, the primary inferred origin in these provinces shifted to the United States. A similar pattern was observed in Xiangkhouang, which borders Vietnam; during the lockdown period, Vietnam and the United Kingdom each accounted for 33% of inferred strain possible origins, but after the lockdown, the dominant origin also shifted to the United States, accounting for 66% (21/32). Notably, introductions from China increased after the lockdown, particularly in the northern provinces such as Luang Namtha, Oudomxay, Luang Prabang, and Houaphan (Figure 7).

**Figure 7.**
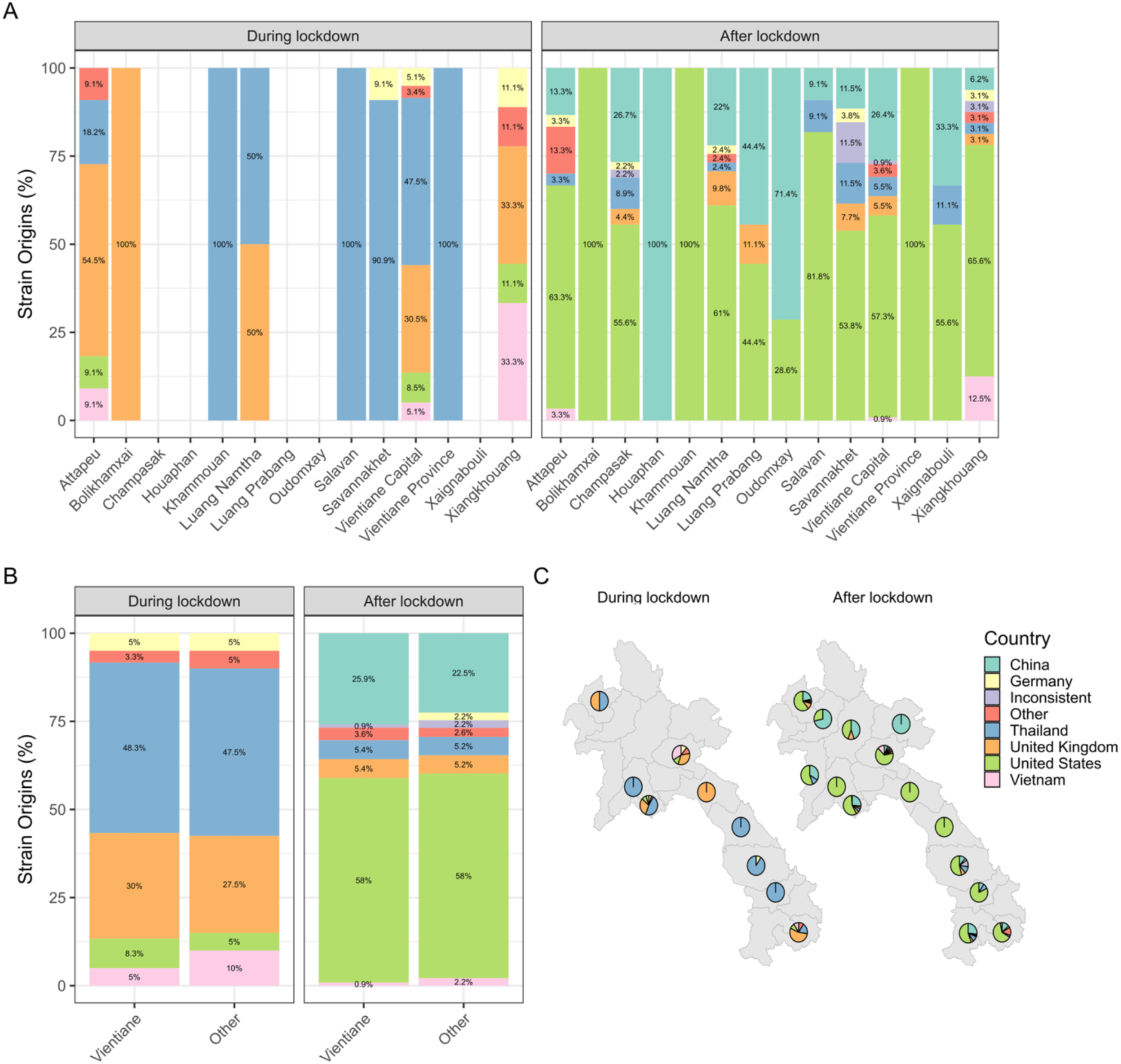
(A) The percentage of strain origins attributed to different countries for each province, separated into during and after lockdown periods. (B) The percentage of strain origins attributed to different countries in Vientiane (combining Vientiane Capital and Vientiane Province) compared to all other provinces across the two periods. (C) Geographic distribution of putative strain origins during and after the lockdown. Mean delay between collection and submission date (days)

To confirm that the geographic origin was not affected by the ancestral state reconstruction methods, we compared results obtained from the DELTRAN and MPPA methods implemented in PastML (Figure S1). Using the MPPA approach gave a higher proportion of introductions which were classed as “uncertain” and a definite geographic origin could not be assigned compared to DELTRAN (17% (17/100) of introductions during the lockdown period and 30.32% (104/343) after the lockdown (Figure S6B)). In contrast, the DELTRAN method provided only 1.75% (6/343) inconclusive results in the post-lockdown analysis (Figure S6A). However, when the inconclusive results were removed, the overall trends remained consistent (Figure S6A and S6C).

We also examined the proportion of samples by country in the phylogenetic tree used for geographic inference, as the inferred country of origin could be biased towards the countries that have the most strains deposited in the public databases. The most represented countries in the tree were the United Kingdom, Germany, the United States, and China, which aligns with the top origin countries inferred in our analysis (Figure S7), with China contributing a significant number of strains only during the later period of the pandemic. However, Thailand and Vietnam comprise only a small fraction of samples in the global tree but account for 48% (48/100) and 7% (7/100) of inferred strain origins during the lockdown period (Figure 6B).

## Discussion

We described the introduction and spread of SARS-CoV-2 variants sampled in Laos during the COVID-19 pandemic. After early success in controlling the virus spread, similar to other Southeast Asian countries, including Vietnam^29^ and Malaysia^30^, we showed that subsequent waves of infection driven by different variants occurred in Laos. We identified a case of the Gamma variant, which has not previously been detected in Laos, but this single strain had no sequenced descendants from any country, suggesting very limited spread of this variant.

The most prevalent lineages across the different waves of COVID-19 in Laos closely match those seen in other countries worldwide, with the first wave caused by the Alpha variant in early 2021 which was rapidly replaced by Delta lineages mid-2021, and subsequent waves of Omicron BA.1, BA.2, and BA.5 variants, and finally the XBB and BA.2.86 descendant lineages which were still circulating at the end of our study period in 2024. Despite the low number of detected cases of the virus until early 2021, we show that two dominant Delta variant lineages, each likely derived from a single introduction, were widely transmitted within Laos after their initial introduction. A review of seven other Southeast Asian nations also identified clusters of high transmission during the Delta variant wave^31^. In contrast, later waves, which generated fewer cases, stemmed from numerous distinct viral introductions, despite travel restrictions still being in place. This may be due to increasing population immunity through vaccination, which began in mid-2021. These large Delta lineage clusters show that a single introduction can seed a large transmission cluster and show the importance of continued surveillance during a pandemic. In contrast, the single strains of the Gamma and Mu variants have no identified descendants within the country. Phylogenetic analyses examining the introduction and proliferation of viral strains can help militaries prepare for subsequent viral pathogens entering congregate settings such as the recruit environment.

Our comparison of viral introductions showed that the proportion of introductions from the Vientiane region and the rest of the country appears similar during a period of travel restrictions and after the restrictions were lifted; however, the number of introductions per sequenced strain, and the probable country of origin of the introductions were different in the two time periods. The early introductions from Vietnam and Thailand may represent cross-border introductions that occurred during a period where air travel was restricted, while the later introductions from the United States and China may represent introductions from air travel, or due to relaxation of outgoing travel restrictions from China. An important consideration for interpretation is that the inference of the country of origin is highly dependent on the deposition of strains into the public sequence databases by national genomic surveillance programmes. The unexpectedly high rate of introductions from the United Kingdom during the lockdown period may represent strains which were not introduced directly to Laos from the United Kingdom, but which were introduced from a third country where the related strains were never sequenced or never deposited into the public databases, and the closest relatives were detected in the United Kingdom due to their extensive genomic surveillance program from early in the pandemic^32^. The lack of information on whether the strains originated in quarantine centres also hampers our interpretation of these results, as we cannot distinguish between the expected detection of SARS-CoV-2 positive patients in quarantine, and introductions which were not detected at border points.

Sampling bias represents a major limitation of our study. Due to low sequencing rates, limited geographical representativeness of the Lao samples analyzed and of the strain sequences available for comparison in the public databases, we may not be able to ascertain the complete picture from genomic surveillance. We detected multiple introductions into the country as first sampled in Xieng Khouang province, but we have a relatively high number of strains collected from this province due to ongoing studies recruiting at Xieng Khouang provincial hospital^13^, while we have only two sequenced strains collected in the neighbouring Houaphan province, which shares a long border with Vietnam. It is also possible that some strains that do not appear to have spread within the country were collected from positive patients admitted to quarantine facilities, as we do not have detailed patient metadata that includes quarantine status. We would expect quarantine facility strains to be less likely to transmit to other patients than those collected outside a quarantine facility, and this may affect the results of our analysis on the introduction characteristics.

The global disparities in SARS-CoV-2 sequencing have been previously investigated^33^, with the recommendation to sequence at least 5% of cases to detect lineages at a prevalence of 0.1 to 1.0%. While the proportion of cases sequenced is an important factor in an effective SARS-CoV- 2 genomic surveillance system, it is also important to consider the number of tests performed and the spatiotemporal bias of the sampling sites^34^. The number of sequenced cases since May 2023 has regularly been greater than the number of reported cases, likely reflecting under-ascertainment of cases once the restrictions were lifted and the number of RT-qPCR tests conducted fell, and the increased use of RDTs which are not reported in national testing statistics. Any future genomic surveillance programme should consider how to maximise the value of whole-genome sequencing in a public health system by choosing a range of sentinel and targeted surveillance sites, and consider that measuring success by proportion of cases sequenced is inappropriate if case numbers are under-reported.

Whole-genome sequencing is a useful tool to study the effectiveness of border control measures during pandemics through the ability to characterise introductory events and inform biosecurity and improve medical readiness for US and other militaries. During lockdown a greater proportion of inferred introductions came from neighbouring countries, which could indicate introductions through informal land borders rather than the formal quarantine system, or introductions through imperfect compliance with border quarantine measures (reported as 73% compliance in a study from Savannakhet Province^35^). If similar lockdown measures were used in future pandemics, improved surveillance could help to improve border surveillance and assess the utility of border control measures. Despite the quarantine measures which reduced the number of viral introductions early in the pandemic, a small number of viral introductions led to a large number of detected cases, demonstrating that border control measures must be used in conjunction with in-country pandemic control measures to effectively suppress viral transmission.

## Disclaimer

Andrew G. Letizia (CAPT, USN, MC) is a military service member. This work was prepared as part of his official duties. Title 17, U.S.C., §105 provides that copyright protection under this title is not available for any work of the U.S. Government. Title 17, U.S.C., §101 defines a U.S. Government work as a work prepared by a military service member or employee of the U.S. Government as part of that person’s official duties. The views expressed in the article are those of the authors and do not necessarily express the official policy and position of the U.S. Navy, the Department of Defense, the U.S. government, or any of the other institutions affiliated with any of the authors.

## Funding and acknowledgements

This work was funded through work unit number ProMIS ID P0153_21_N2 as a component of the Armed Forces Health Surveillance Branch (AFHSB) Global Emerging Infections Surveillance program (GEIS). Additionally, this work was supported by grants from Wellcome (222574/Z/21/Z and 220211/Z/20/Z). We thank all participants to the case-based surveillance study as well as all staff who enrolled patients during the pandemic and processed samples. For the purpose of Open Access, the author has applied a CC BY public copyright licence to any Author Accepted Manuscript version arising from this submission.

## Data availability

Viral sequences are available on GISAID as EPI_SET_251209fy. Supplementary Table 2 lists the sequences used in this study with GISAID identifiers, and Genbank accession numbers where available.

## Supplementary Figures

**Figure S1.**
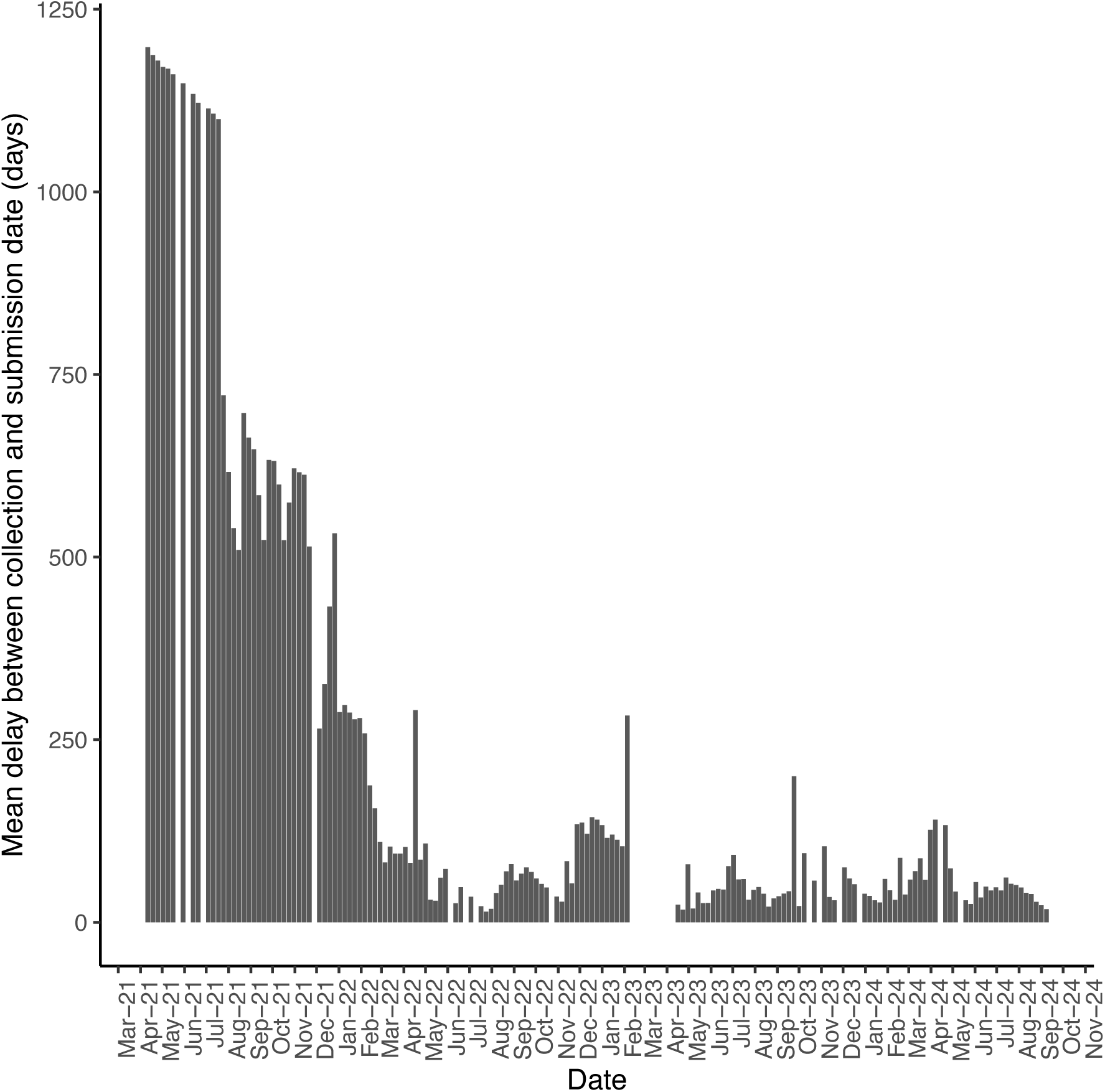
A plot of the mean weekly time delay between sample collection (‘Collection Date’) and submission (‘Submission Date’) of the sequence data to GISAID.

**Figure S2.**
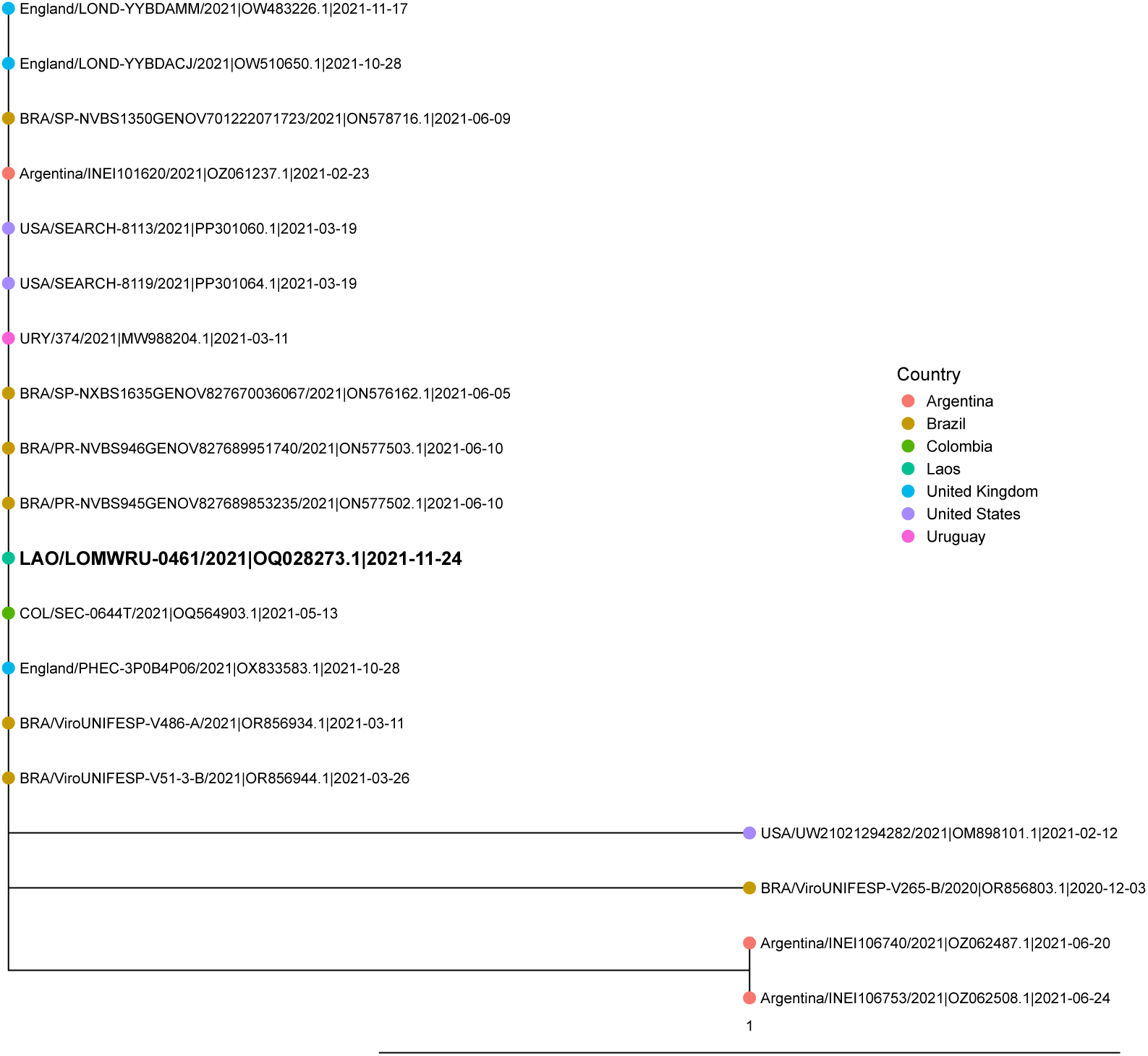
A phylogenetic tree showing Laos strain LOMWRU-0461 and the 20 most closely related to this strain taken from UShER, showing the number of mutations from the Wuhan reference strain to each individual strain. The coloured tip labels give the country for each strain. The scale bar shows the number of mutations.

**Figure S3.**
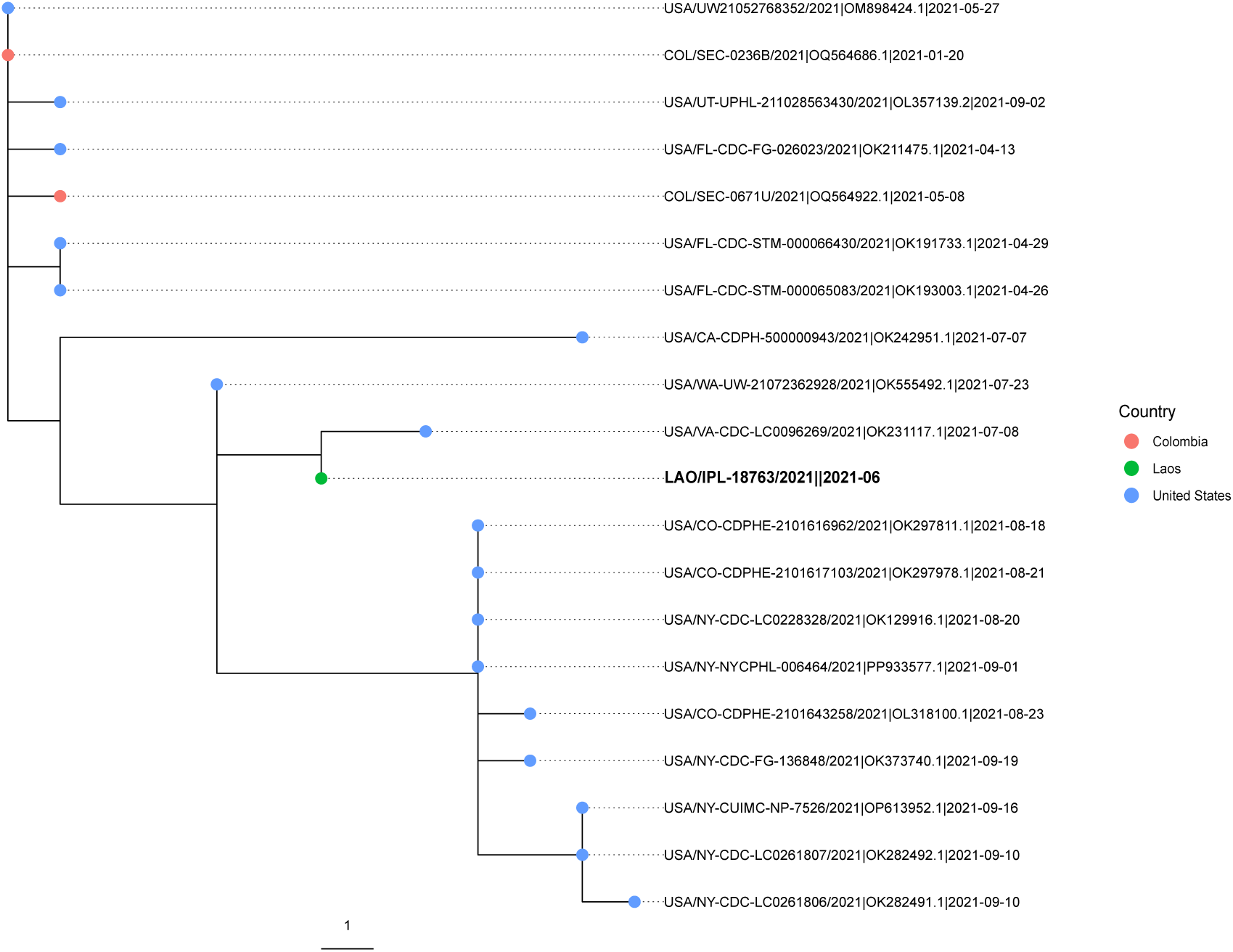
A phylogenetic tree showing Laos strain IPL-18763 and the 20 closest neighbours to this strain taken from UShER, with a scale bar showing the number of mutations from the Wuhan reference strain to each individual strain. The coloured tip labels give the country for each strain. The scale bar shows the number of mutations.

**Figure S4.**
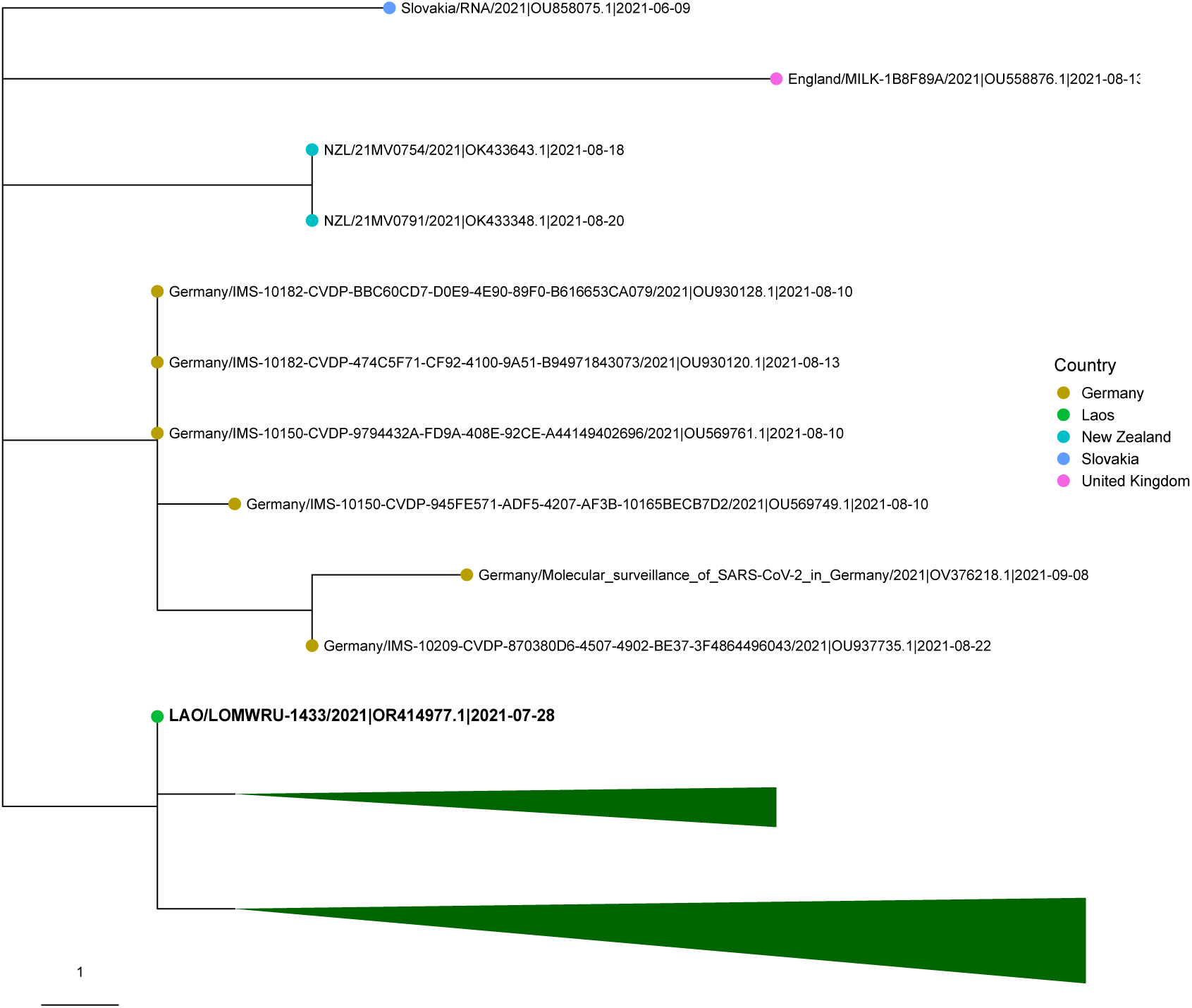
A phylogenetic tree showing Laos strain LOMWRU-1433 and the 643 closest neighbours to this strain taken from UShER, showing the number of mutations from the Wuhan reference strain to each individual strain. There are 633 strains from Laos, which have been collapsed into two green triangles representing 29 and 604 strains, respectively. The remaining 10 strains are displayed individually. The tip colours give the country of origin for each strain. The scale bar shows the number of mutations.

**Figure S5.**
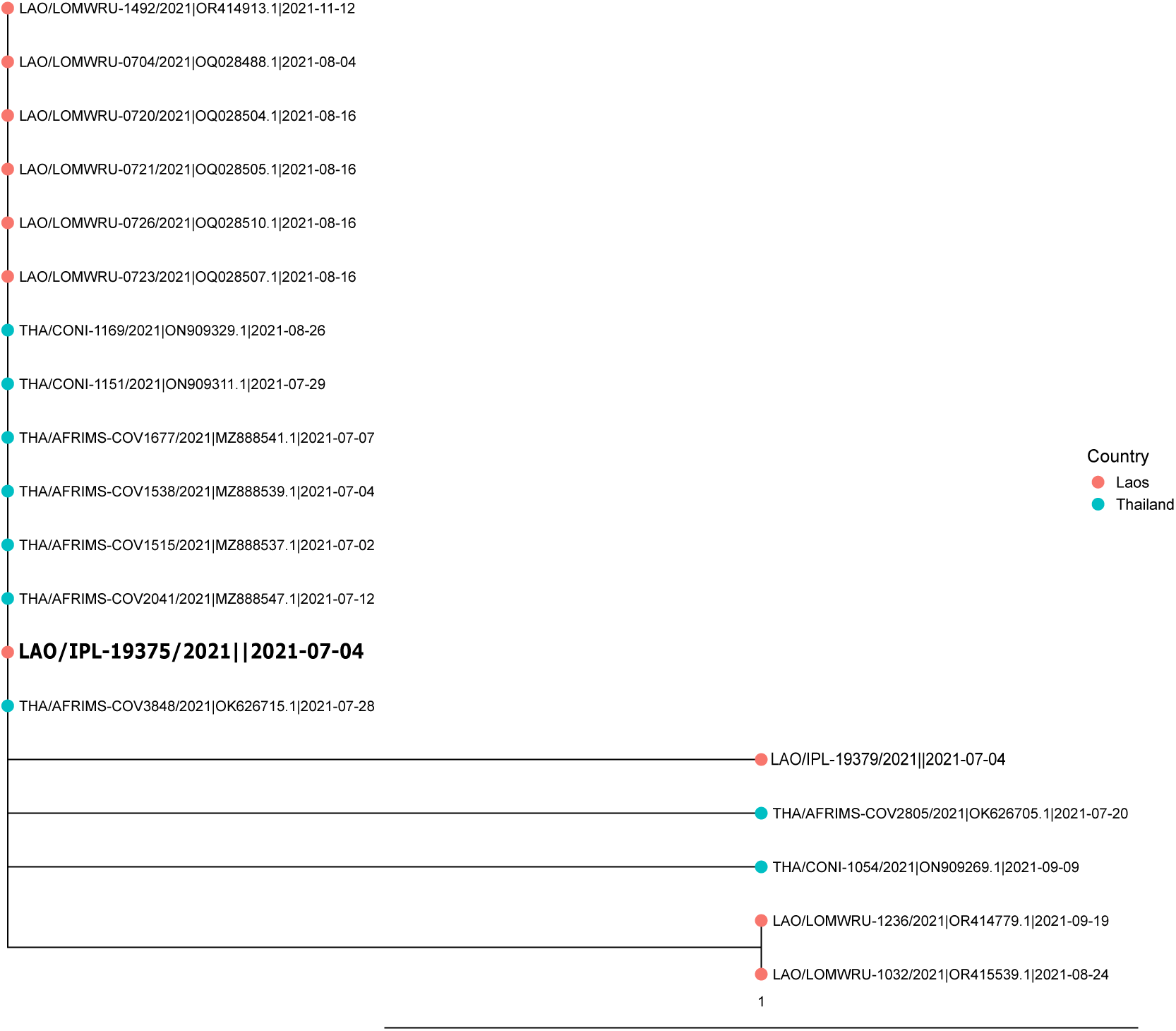
A phylogenetic tree showing Laos strain IPL-19375 and the 20 closest neighbours to this strain taken from UShER, showing the number of mutations from the Wuhan reference strain to each individual strain. The branch and tip colours give the country of origin for each strain. The scale bar shows the number of mutations.

**Figure S6.**
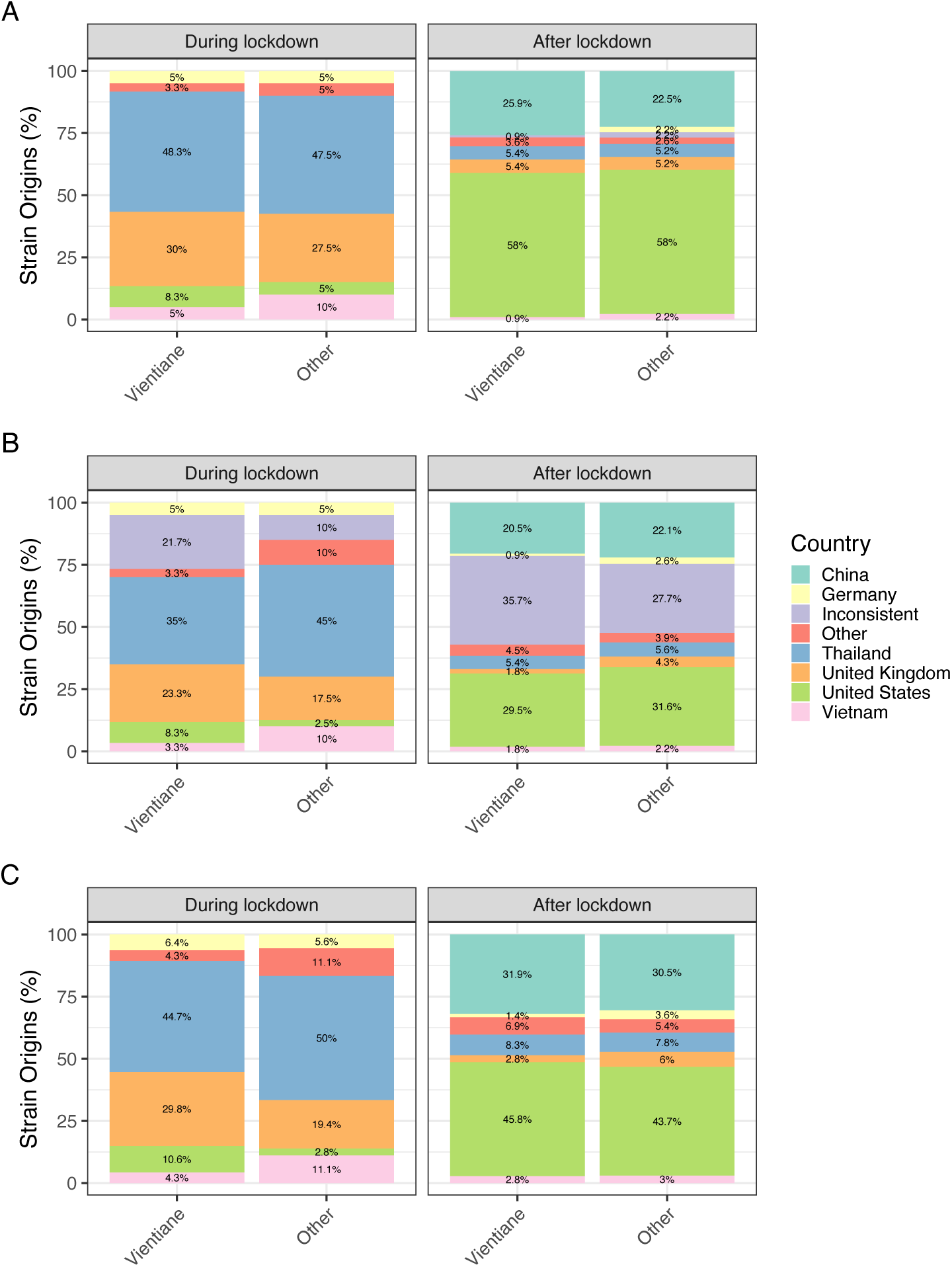
Percentage of strains with a probable origin per country for Vientiane (combining Vientiane Capital and Vientiane Province) compared to all other provinces across the two periods using PastML using DELTRAN and MPPA methods. (A) The same comparison using PastML with DELTRAN method. (B) PastML with MPPA method. (C) The plots from panel A using PastML with MPPA method but removing inconsistent results.

**Figure S7.**
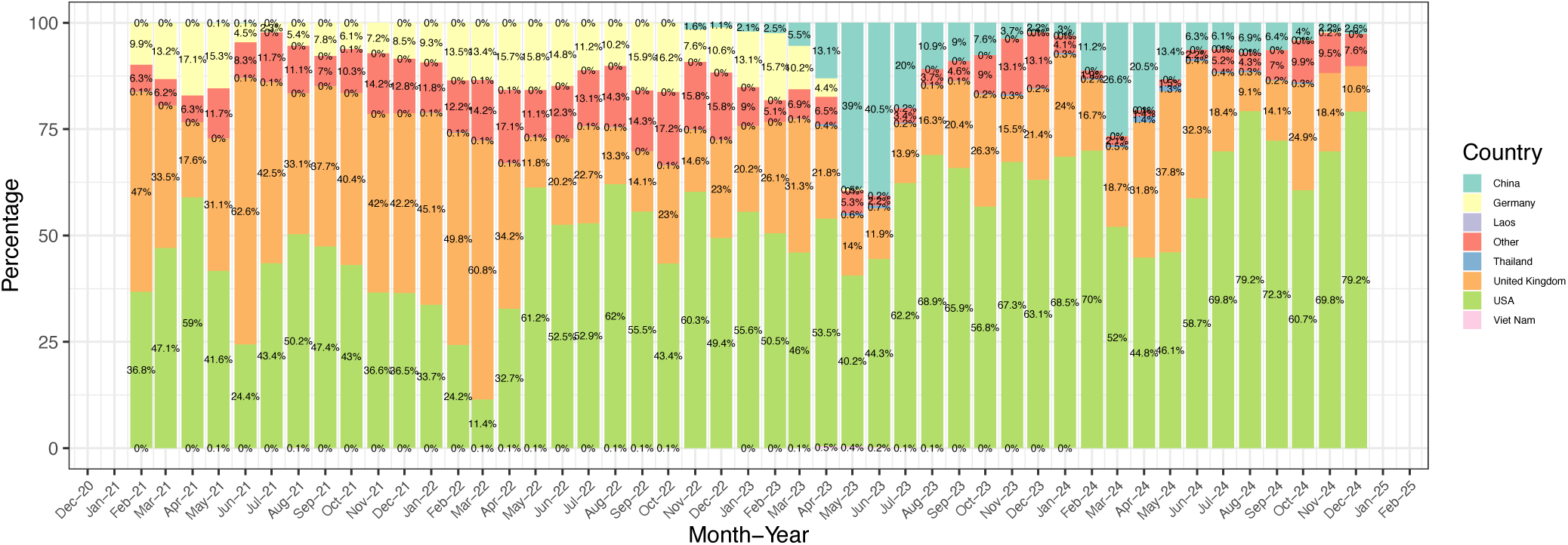
Monthly distribution of sequenced sample by country in the global phylogenetic tree used for inferefence of introduction origins. The stacked bar chart displays the percentage of samples contributed by each country over time.

**Table S1.** Total number of SARS-CoV-2 PCR positive samples, the number where sequencing was attempted, and the total number of samples submitted to GISAID, for the isolates collected at the Lao-Oxford-Mahosot Wellcome Trust Research Unit.

| Province | Number of SARS-CoV-2 positive samples by RT-qPCR | Number of samples where SARS-CoV-2 sequencing was attempted | Number of sequences submitted to GISAID |
| --- | --- | --- | --- |
| Vientiane Capital | 3045 | 1208 | 1106 |
| Phongsali | 1 | 0 | 0 |
| Luang Namtha | 373 | 187 | 147 |
| Oudomxay | 3 | 1 | 1 |
| Bokeo | 3 | 1 | 1 |
| Luang Prabang | 5 | 1 | 1 |
| Houaphan | 3 | 0 | 0 |
| Xaignabouli | 9 | 1 | 1 |
| Xieng Khouang | 255 | 251 | 201 |
| Vientiane | 94 | 47 | 46 |
| Bolikhamxai | 16 | 3 | 2 |
| Khammouan | 113 | 90 | 84 |
| Savannakhet | 62 | 22 | 19 |
| Salavan | 57 | 25 | 17 |
| Sekong | 0 | 0 | 0 |
| Champasak | 2 | 1 | 0 |
| Attapeu | 234 | 210 | 172 |
| Xaisomboun | 24 | 11 | 11 |
| Unknown province | 1224 | 243 | 178 |
| Total | 5523 | 2302 | 1987 |

